# A Qualitative Study of Patient and Healthcare Provider Perspectives on Mobile Health Assessments for Cervical Spondylotic Myelopathy

**DOI:** 10.64898/2026.03.04.26347622

**Authors:** Pranay Singh, Sriharsha Gonuguntla, Erdong Chen, Aryan Pradhan, Isaac Becker, Nathan Xu, Benjamin Steel, Faraz Arkam, Salim Yakdan, Haris Naveed, Zhuoran Wang, Wenwen Guo, Zachary Wilt, Jetan Badhiwala, Daniel Hafez, John Ogunlade, Wilson Z. Ray, Zoher Ghogawala, Caitlin Kelleher, Jacob Greenberg

**Affiliations:** Department of Neurosurgery, Washington University in St. Louis School of Medicine, St. Louis, Missouri, USA; University of South Florida Morsani College of Medicine, Tampa, Florida, USA; Saint Louis University School of Medicine, St. Louis, Missouri, USA; Rothman Orthopaedics, Philadelphia, Pennsylvania, USA; Department of Surgery, University of Toronto, Toronto, Ontario, Canada; Department of Surgery, UMass Chan Medical School, Burlington, Massachusetts, USA; Department of Computer Science and Engineering, Washington University in St. Louis, St. Louis, Missouri, USA

**Author notes:** Corresponding Author: Jacob Greenberg MD MSCI, Name: Jacob Greenberg, Current Institution: Washington University in St. Louis School of Medicine. Co-first authors.

**Keywords:** cervical spondylotic myelopathy, mobile health, digital biomarkers, qualitative research, stakeholder perspectives, smartphone assessment

## Abstract

**Objective:** Evaluating and monitoring patients with cervical spondylotic myelopathy (CSM) remains a challenge due to limited tools for assessing objective neurological disability longitudinally and in the home environment. Given their prevalence and low cost, mobile health (mHealth), and specifically smartphone technologies offer a promising approach to fill this gap. This study explored stakeholder perspectives on the role of mHealth in CSM monitoring to inform development of a smartphone-based assessment application.

**Methods:** We conducted semi-structured interviews with 15 patients with CSM and 14 healthcare providers (spine surgeons, physical therapists, and occupational therapists). Interviews explored current assessment practices, perceived limitations, and attitudes toward mHealth integration. Data were analyzed using thematic analysis.

**Results:** Two major themes emerged from provider interviews: (1) diagnosing and monitoring CSM is challenging due to limitations in current tools, and (2) mHealth presents significant opportunities but requires thoughtful integration. Providers described current methods and technologies, clinical signs and symptoms, and challenges evaluating patients. Current tools were viewed as inadequate for precision medicine, with inter-rater variability and inability to capture real-world function. Within the second theme, providers identified ways mHealth could improve care, challenges for integration, and practical implementation considerations. Patients expressed strong interest in objective, longitudinal monitoring of gait, dexterity, and daily function.

**Conclusions:** Stakeholders recognized substantial potential for mHealth to address unmet needs in CSM assessment. Successful implementation will require intuitive design, electronic medical record integration, and attention to accessibility. These findings provide a foundation for user-centered development of digital health tools in CSM care.

## Introduction

Cervical spondylotic myelopathy (CSM) is the most common cause of spinal cord dysfunction in older adults worldwide.^1^ Progressive spondylotic degeneration and compression of the cervical spinal cord lead to upper motor neuron signs, gait disturbance, and loss of fine motor control, severely impairing quality of life.^2^ Despite its prevalence and clinical significance, timely diagnosis and quantitative monitoring of progression remain challenging.

Current clinical assessments have substantial limitations. Most monitoring relies on patient-reported tools, including the modified Japanese Orthopaedic Association (mJOA) scale^3^ and Neck Disability Index.^4^ However, such approaches fail to quantify objective neurological abilities relevant to CSM and demonstrate limited sensitivity to subtle neurological changes.^5,6^ Consequently, there is a critical need for accessible, objective, and reproducible markers of neurological function in CSM, particularly for longitudinal, real-world use.

Mobile health (mHealth) technologies represent a promising solution. Smartphones and wearable devices can capture sensor-based digital biomarkers that objectively quantify neurologic function. Such tools have demonstrated feasibility in Parkinson’s disease (PD), multiple sclerosis (MS), and traumatic brain injury.^7–10^ However, within neurosurgery, and particularly in the management of CSM, clinical use of mHealth remain limited.^11–15^

Lessons from other fields suggest that understanding end-user perspectives is essential for mHealth development.^16–18^ For example, while early stakeholder involvement is considered paramount,^19,20^ only 19% of smartphone studies in PD formally evaluated usability,^19^ and adherence remains inconsistent.^21^ Therefore, understanding how patients and healthcare professionals perceive mHealth feasibility, utility, and barriers early in development is critical.

Recently, we developed SynapTrack, a smartphone application using built-in sensors to provide objective measures of disability in CSM.^22^ During early development, we conducted semi-structured interviews with clinicians and patients to guide development and understand current practices, perceptions, and unmet needs. This study presents stakeholder perspectives on the potential clinical role, challenges, and implementation considerations relevant to smartphone assessments for CSM.

## Methods

### Study Design and Ethics

Semi-structured interviews explored stakeholder perspectives on smartphone-based monitoring for CSM. The study received ethical approval from the Washington University in St. Louis Institutional Review Board. All participants provided informed consent.

### Participants

We recruited patients with CSM and healthcare providers who regularly care for this population. Patients diagnosed with CSM were recruited from the neurosurgery department of Washington University in St. Louis School of Medicine. Patients with other neurodegenerative diseases (e.g., amyotrophic lateral sclerosis, PD) and those with non-cervical spine surgery within the preceding two months were excluded.

Healthcare providers who regularly care for patients with CSM were recruited from the same academic medical center as well as other health systems and private practices. The provider sample included spine surgeons (neurosurgery and orthopedic surgery), physical therapists, and occupational therapists, recruited through professional networks among the research team.

### Data Collection

Patient interviews were conducted between August and November 2024 in a clinical laboratory environment following structured testing with an early iteration of SynapTrack. Provider interviews were conducted via video conference. All interviews were recorded and professionally transcribed (Landmark Associates, Phoenix, AZ).^23^ Semi-structured interview guides were developed separately for patients and providers (Table 1). Guides promoted open-ended discussion while capturing perspectives on several core concepts. Patient interviews (∼10 minutes) explored symptoms, self-monitoring, and SynapTrack usability. Provider interviews explored assessment methods, perceived limitations, and perspectives on mHealth.

**Table 1.** Semi-Structured Interview Topics for Patients and Healthcare Providers.

| Patient Interview Topics | Provider Interview Topics |
| --- | --- |
| Confidence in task completion | Professional background and patient population |
| Concerns about task completion | Tools currently used to assess patients with CSM |
| Addressing difficulties during challenging tasks | Use of technology in clinical workflow |
| Motivation to continue using the app | Information about CSM patients difficult to obtain |
| Incorporating app use into daily life | Challenges in assessing CSM patients |
| Engagement with other health apps | Potential roles for digital health applications |
| Symptoms of cervical myelopathy | Tasks/activities for a patient-facing digital app |
| Monitoring myelopathy severity | Preferred methods of accessing digital data |
| Overall experience with the app | Preferred frequency of digital assessments |
|  | Additional feedback and recommendations |

*CSM = Cervical Spondylotic Myelopathy.*

**Table 2.** Patient Demographics (N = 15)

| **Characteristic** | **n (%)** |
| --- | --- |
| Sex, Male | 7 (46.7) |
| Age, years, mean (SD) | 61.7 (10.1) |
| **Race** |  |
| White | 12 (80.0) |
| Black | 3 (20.0) |
| **Education (n = 14)** |  |
| High school diploma | 5 (35.7) |
| College degree | 6 (42.9) |
| Graduate/professional degree | 3 (21.4) |
| **Employment** |  |
| Working | 5 (33.3) |
| Retired | 6 (40.0) |
| On disability | 4 (26.7) |
| **Area of Residence** |  |
| Metropolitan | 14 (93.3) |
| Rural | 1 (6.7) |
| mJOA Score, mean (SD) (n = 14) | 14.1 (2.7) |

*SD = Standard Deviation; mJOA = modified Japanese Orthopaedic Association scale.*

### Data Analysis

Patient and provider interview data were analyzed separately using thematic analysis following Braun and Clarke’s six-step framework^24–26^: (1) familiarization through thorough reading; (2) systematic coding; (3) organization into potential themes; (4) theme review and refinement; (5) defining themes; and (6) final reporting. Two researchers analyzed each dataset, discussing categorizations at each step to reach consensus. A third, or if needed fourth, researcher then reviewed and helped revise the themes to reach the final scheme.

### Validation

Thematic categorizations were validated separately for each dataset. Initial themes were reviewed by clinical and human-computer interaction experts. A blinded researcher then independently assigned each quote to established categories (Figure 1). Percent agreement and Cohen’s kappa were calculated. For provider interviews, a second validation was conducted after refining category descriptions. Initial validation revealed frequent misclassification between two conceptually overlapping categories which were subsequently consolidated, reducing the total from seven to six categories.

**Figure 1.**
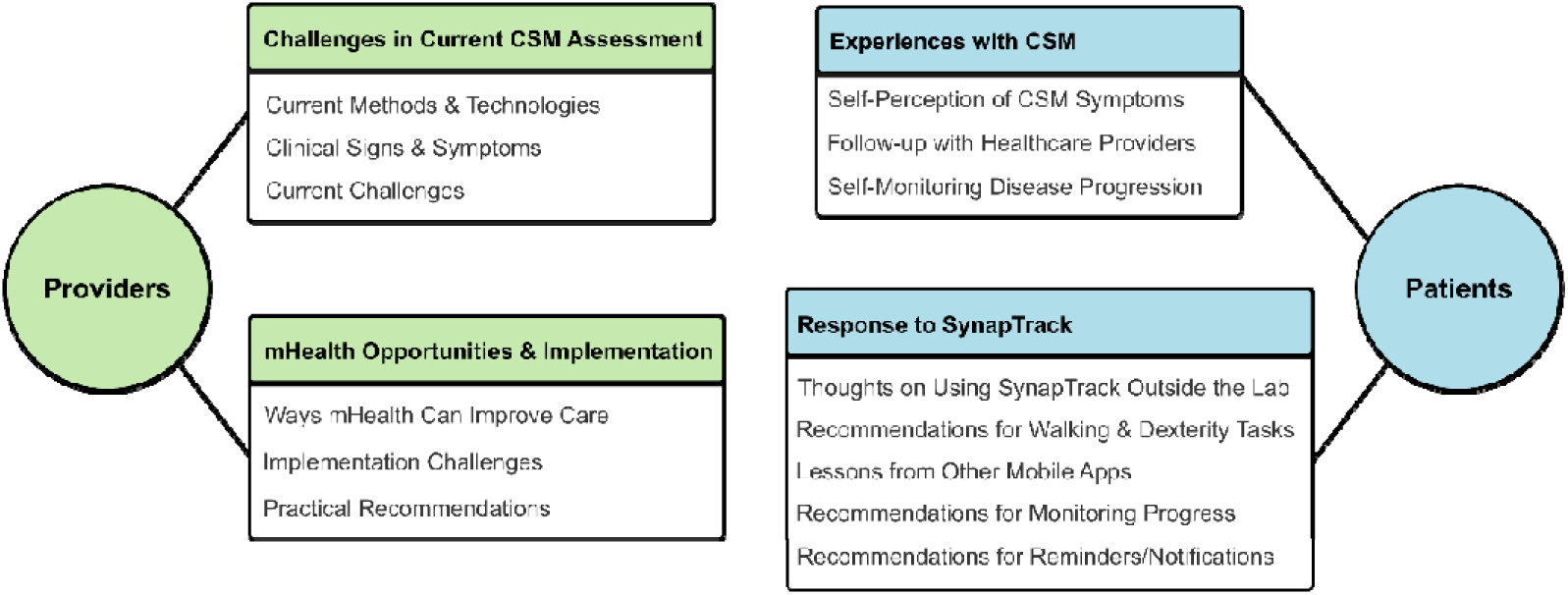
Summary of thematic analysis from semi-structured interviews with healthcare providers (n=14) and patients (n=15) regarding mHealth for DCM. Provider themes included current assessment challenges (blue) and mHealth opportunities and implementation considerations (purple). Patient categories encompassed experiences with DCM and responses to SynapTrack (green). DCM = degenerative cervical myelopathy; mHealth = mobile health.

## Results

### Provider Themes

#### Participant Characteristics

14 healthcare providers completed semi-structured interviews. Provider characteristics are summarized in Table 3 (demographics available for 13/14). The sample included 4 orthopedic surgeons, 5 physical therapists, 3 neurosurgeons, and 2 occupational therapists across 8 institutions. Most practiced in academic or hospital settings (69.2%). Median years in practice was 5 (range 3–43), and median CSM patients seen per year was 25 (range 0–200). Provider data were organized into two themes (Tables 4–5, Figure 1).

**Table 3.** Healthcare Provider Demographics (N = 13)

| **Characteristic** | **n (%)** |
| --- | --- |
| **Occupation** |  |
| Orthopedic surgeon | 4 (30.8) |
| Physical therapist | 5 (38.5) |
| Neurosurgeon | 2 (15.4) |
| Occupational therapist | 2 (15.4) |
| **Practice Setting** |  |
| Academic/hospital | 9 (69.2) |
| Private clinic | 2 (15.4) |
| Outpatient rehabilitation | 2 (15.4) |
| **Gender** |  |
| Male | 7 (53.8) |
| Female | 6 (46.2) |
| Age, years, mean (range) | 39.2 (26–77) |
| Years in practice, median (range) | 5 (3–43) |
| CSM patients/year, median (range) | 25 (0–200) |

*CSM = Cervical Spondylotic Myelopathy.*

**Table 4.** Healthcare Provider Theme 1: Diagnosing and Monitoring CSM Is Challenging Due to Limitations in Current Tools.

| Category | Representative Quotes |
| --- | --- |
| **Current Methods and Technologies Used to Assess Patients with CSM** | “Many of us think that the mJOA is a relatively bad assessment for [myelopathy]. (Neurosurgeon).<br>“Love them, but...we want precision medicine.” (Orthopedic surgeon)<br>“We’re just still sticking to the tried and true like goniometry, paper documentation. In our world, we don’t seem to have a lot of new technology.” (Occupational therapist)<br>“I don’t do anything with wearables... but it seems like a good way to collect data from people.” (Neurosurgeon) |
| **Clinical Signs and Symptoms of CSM** | “I ask about dropping objects, clumsiness with their hands, difficulty with jewelry and small objects.” (Orthopedic surgeon)<br>“Pain—which is not a very sensitive sign of myelopathy—is also on the top of the list.” (Neurosurgeon)<br>“Bladder function is changing, unspecific but it’s still good in the context of numbness and walking and hand function.” (Neurosurgeon)<br>“I think spasticity would be pretty difficult to measure.” (Physical therapist) |
| **Current Challenges Evaluating Patients with CSM** | “I think what patients are doing at home is one of the biggest question marks.” (Physical therapist)<br>“The static assessment of their function on one specific day... [is] of limited usefulness.” (Neurosurgeon).<br>“Patients have, on average, pretty poor recall. If we can collect it in real time, I think it would be better.” (Orthopedic surgeon)<br>“These large clinics often have trainees with me... there’s variability. What one person’s assessment of strength or reflex or coordination is maybe different than somebody else.” (Orthopedic surgeon)<br>“There isn’t always a perfect correlation between clinical symptoms and imaging findings.” (Orthopedic surgeon)<br>“What I’m looking for would be a reproducible standardized measure of clinical function that correlates to patient outcomes. So if I had a tool that would reliably tell me... exactly where they stood along the disease severity, and I could monitor either any change for the better or worse, and it was reproducible and accurate. It would be a very helpful tool for me.” (Orthopedic surgeon). |

*CSM = Cervical Spondylotic Myelopathy; mJOA = modified Japanese Orthopaedic Association scale.*

**Table 5.** Healthcare Provider Theme 2: mHealth Presents Significant Opportunities but Requires Thoughtful Integration.

| Category | Representative Quotes |
| --- | --- |
| **Ways Mobile Health Can Improve Care for Patients with CSM** | “The focus should be on the things are a bit controversial. The one that are borderline, you're not sure based on imaging or clinical finds. That's the group that will benefit, both patient and surgeon.” (Neurosurgeon)<br>“That would help push me to offer somebody surgery or not, which is a big deal.” (Neurosurgeon)<br>“Mapping cervical myelopathy would be analogous to looking at hemoglobin A1C trends.” (Orthopedic surgeon)<br>“It helps empower the patients... and helps surgeons reassure them and assuage fears.” (Orthopedic surgeon)<br>“It would make our workflow a lot easier if it just spat out the information for us.” (Occupational therapist) |
| **Challenges Associated with and Advice for Integrating Mobile Assessments into CSM Care** | “If their dexterity is bad because they're myelopathic, they're less likely to enter the data in a phone properly.” (Orthopedic surgeon)<br>“There could be a lot of barriers for some of these patients...from different socioeconomic backgrounds.” (Physical therapist)<br>“[If] it's not part of the electronic medical record...[it's] just kind of fragmented.” (Physical therapist)<br>“If it's not super seamless...unless I'm really getting a lot of benefit in terms of conversation and the way it's influencing my care, I think it's something that would be easy to say I'm just not gonna use this with this patient.” (Occupational therapist)<br>“Finding a signal in the data that's meaningful is probably going to be challenging.” (Neurosurgeon) |
| **Feedback Related to Practical Implementation of mHealth in Clinical Practice** | “It should be more frequent at the beginning, within the first six months maybe every month...then after six months...every three or six months.” (Neurosurgeon)<br>“I love when things are integrated into an EMR... that would be gold standard.” (Occupational therapist)<br>“Have it in digital data, which would be great, bar graphs, charts, that kinda thing so we're not having to flip through countless amounts of paperwork. Just have a quick comparative first session, today's session.” (Occupational therapist)<br>“Obviously there's the issue of alarm fatigue.” (Neurosurgeon).<br>“If the app was notifying the patient...‘Hey, the trend is trending in the wrong direction. You should make an appointment with your provider.’ I think that would be pretty cool.” (Orthopedic surgeon) |

*EMR = Electronic Medical Record; mHealth = Mobile Health; CSM = Cervical Spondylotic Myelopathy.*

### Theme 1: Diagnosing and Monitoring CSM Is Challenging Due to Limitations in Current Tools

Three categories emerged: (1) current methods and technologies used to assess patients with CSM, (2) clinical signs and symptoms of CSM, and (3) current challenges evaluating patients with CSM (Table 4).

***Current Methods and Technologies.*** Clinicians primarily relied on traditional clinical evaluations including history, neurological examination (including assessment of gait, hand dexterity, balance, and reflexes), and imaging (computed tomography and magnetic resonance imaging). Objective bedside measures, such as grip dynamometry, were occasionally incorporated:

> *“Other than clinic exams? No…the only distinguishing characteristic in my visit was that I would use a grip dynamometer…in addition to their subjective complaints.” (Orthopedic surgeon)*.

Standardized questionnaires were commonly used to quantify functional limitations, including the Patient-Reported Outcomes Measurement Information System (PROMIS),^27^ mJOA scale,^3^ Neck Disability Index,^4^ and Canadian Occupational Performance Measure.^28^ However, providers expressed significant dissatisfaction with these tools. One surgeon noted:

> *“Many of us think that the mJOA is a relatively bad assessment for [myelopathy].” (Neurosurgeon)*.

Another described current categorical measures as fundamentally inadequate for individualized care:

> *“…we want precision medicine. If we can turn those categories into a continuous measure… it would help you assist with your decision making.” (Orthopedic surgeon)*

Use of digital or mHealth tools was limited and varied widely among clinicians. Physical and occupational therapists reported employing patients’ wearable devices for activity monitoring in rehabilitation contexts. However, most participants reported limited or no use:

> *“I don’t do anything with wearables… but it seems like a good way to collect data from people.” (Neurosurgeon)*
>
> *“We’re just still sticking to the tried and true like goniometry, paper documentation.” (Occupational therapist)*

***Clinical Signs and Symptoms*.** Providers described CSM presentations including gait disturbances (slowed speed, shorter strides), fine motor dysfunction, and sensory changes such as numbness and tingling. One provider noted:

> *“I ask about dropping objects, clumsiness with their hands, difficulty with jewelry and small objects.” (Orthopedic surgeon)*

Pain and urinary symptoms were viewed as nonspecific, while upper motor neuron signs were consistently identified on examination.

***Current Challenges Evaluating Patients with CSM*.** Healthcare professionals described persistent challenges in capturing an accurate and comprehensive picture of patients’ functional abilities. Therapists noted that clinic-based assessments provided only a limited view:

> *“I think that’s a constant question mark of, how can we improve understanding of real-world activity performance over, what does their capacity look like in the clinic, ‘cause we really just get this small window into their life.” (Physical therapist)*

Subtle functional changes, such as declining handwriting quality or increasing frequency of dropped objects, were reported to go unnoticed or described as inconsistent due to recall bias and subjectivity:

> *“Oftentimes patients…don’t accurately assess their own level of deficit.”(Occupational therapist)*
>
> *“Patients have, on average, pretty poor recall. If we can collect it in real time, I think it would be better.” (Occupational therapist)*

Variability in how in-person assessments are conducted and interpreted was reported to pose another significant hurdle:

> *“It’s not always the same person conducting the examination. So there’s variability. What one person’s assessment of strength or reflex or coordination is maybe different than somebody else.” (Orthopedic surgeon)*.
>
> *“…current practice has not been very well standardized, which [makes it] difficult to interpret from one assessor to another.” (Orthopedic surgeon)*.

Surgeons particularly struggled with borderline cases, where clinical decision-making is most difficult:

> *“If I see a patient with mild myelopathy, it’s hard to counsel them on their risk of progression.” (Orthopedic surgeon)*
>
> *“The main issue is just essentially verifying the truth of what is going on clinically with your patient. You sort of have to just figure it out yourself… and there has to be a better way to do that.” (Neurosurgeon)*

The imperfect correlation between MRI findings and clinical symptoms was described as another challenge. One surgeon highlighted the absence of standardized, objective methods for quantifying CSM severity and tracking progression:

> *“What I’m looking for would be a reproducible standardized measure of clinical function that correlates to patient outcomes. So if I had a tool that would reliably tell me… exactly where they stood along the disease severity, and I could monitor either any change for the better of worse, and it was reproducible and accurate. It would be a very helpful tool for me.” (Orthopedic surgeon)*

### Theme 2: mHealth Presents Significant Opportunities but Requires Thoughtful Integration

Three categories emerged: (1) ways mobile health can improve care for patients with CSM, (2) challenges associated with and advice for integrating mobile assessments into CSM care, and (3) feedback related to practical implementation of mHealth in clinical practice (Table 5).

***Ways Mobile Health Can Improve Care for Patients with CSM.*** Across interviews, there was strong consensus that mHealth tools could bridge gaps in objective assessment and enable monitoring of symptoms outside the clinic. Providers noted that such data could clarify disease trajectories, distinguish stable mild from progressive cases, and improve intervention timing. Surgeons emphasized that borderline cases would benefit most:

> *“The focus should be on the things [that] are a bit controversial. The one[s] that are borderline, you’re not sure based on imaging or clinical finds. That’s the group that will benefit, both patient and surgeon.” (Neurosurgeon)*

Surgical decision-making was identified as a key application:

> *“That would help push me to offer somebody surgery or not, which is a big deal… what we’re always looking for is, when do you know when it’s time to do surgery on somebody?” (Neurosurgeon)*

Tools capturing real-time metrics of gait, dexterity, or balance were viewed as enabling more personalized, precision-based care:

> *“A better system for understanding disease severity would be very good in terms of gait disturbance…and fine motor tasks.” (Orthopedic surgeon)*

Therapists reported that home-based data could streamline clinical encounters by reducing reliance on patient recall:

> *“That would be wonderful if we could get a bird’s eye view on what’s happening at home versus what they’re telling us…being like, ‘Remember the other day, you struggled with X, Y, and Z in the home.’” (Occupational therapist)*

One surgeon compared the value of longitudinal mHealth tracking to established chronic disease monitoring:

> *“If a patient’s able to collect that data, and then, show me either graphically, or numerically, I guess, in my mind, mapping cervical myelopathy would be analogous to looking at hemoglobin A1C trends.” (Orthopedic surgeon)*

Participants also described mHealth as strengthening patient engagement and providing psychological benefits:

> *“Post-operative patients would always call saying ‘I’m getting worse’…Very rarely were they actually getting worse…giving them a way to test themselves and measure their own recovery and ensure that they’re not getting worse…I think that’ll be really, really helpful.” (Orthopedic surgeon)*
>
> *“It’s important to help the patient feel they’re being followed…it reduces their anxiety.” (Neurosurgeon)*
>
> *“It helps empower the patients and understand the decisions they’re making.” (Orthopedic surgeon)*

Workflow efficiency was another anticipated benefit:

> *“Oh, my gosh, it would make our speed so much easier, our efficiency so much easier and our documentation. It would make our workflow a lot easier if it just spat out the information for us.” (Occupational therapist)*

***Challenges Associated with and Advice for Integrating Mobile Assessments into CSM Care.*** Participants identified multiple barriers to implementing mobile assessments. A primary concern was technical usability, particularly given that many CSM patients experience fine motor impairment and may be less familiar with smartphone technology:

> *“If their dexterity is bad because they’re myelopathic, they’re less likely to enter the data in a phone properly.” (Orthopedic surgeon)*

Equity and accessibility were emphasized as important considerations:

> *“Maybe they’re homeless, or maybe they live in areas that are not safe to walk around, or don’t have good access to public transportation, or even caregivers who can take care of them. I feel like that can be a huge barrier… reaching their fullest potential. I wonder how we can leverage an app like this… or make sure when we’re designing it to perhaps overcome some of those barriers.” (Physical therapist)*

Ease of use for both patients and providers was described as essential for sustained adoption:

> *“If it’s not super seamless and quick…unless I’m really getting a lot of benefit, it’s something I would stop using.” (Occupational therapist)*

Providers expressed significant concern about workflow integration. Many cited limited time to review digital data:

> *“Into clinical workflow, I think the biggest piece is just we are already pressed for time oftentimes in the outpatient clinic.” (Occupational therapist)*

Privacy and institutional constraints were viewed as additional obstacles:

> *“Always been an issue with patient confidentiality. So how the data is being used, where it’s being stored, who has access to it.” (Orthopedic surgeon)*

One surgeon recognized the challenge of translating raw digital data into actionable insights:

> *“Finding a signal in the data that’s meaningful is probably going to be challenging.” (Neurosurgeon)*

***Feedback Related to Practical Implementation of mHealth in Clinical Practice.*** Participants provided specific recommendations on implementation parameters. Assessment frequency recommendations ranged from every two weeks to every six months, with consensus that cadence should align with clinical phase and patient-specific factors:

> *“It’s better to be more frequent at the beginning, within the first six months maybe every month…then after six months maybe every—it’s an app, if it’s simple, every three months or every six months…[if] I feel my symptom are changing, then you can just test yourself, as a add-on.” (Neurosurgeon)*

A strong theme emerged around the need for integration with electronic medical records (EMRs):

> *“Certainly, I would not be opposed to looking at it in an app, but I love when things are integrated into an EMR… that would be gold standard.” (Occupational therapist)*

Clinicians preferred viewing data directly within patient charts, via visual displays such as line graphs, bar charts, or scatter plots:

> *“Have it in digital data, which would be great, bar graphs, charts, that kinda thing so we’re not having to flip through countless amounts of paperwork. Just have a quick comparative first session, today’s session.” (Occupational therapist)*

Respondents expressed mixed views on provider-facing alerts. While some valued notifications that patients had completed assessments, others cautioned about clinician burden:

> *“Obviously there’s the issue of alarm fatigue. You know, for clinicians and anything related to the EHR.” (Neurosurgeon)*

Patient-facing alerts for concerning trends were generally viewed favorably:

> *“If the app was notifying the patient…‘Hey, the trend is trending in the wrong direction. You should make an appointment with your provider.’ I think that would be pretty cool.” (Orthopedic surgeon)*

### Patient Themes

#### Patient Characteristics

15 patients completed semi-structured interviews. Patient characteristics are summarized in Table 2. Mean age was 61.7 years (SD 10.1), 46.7% were male, 80% identified as White, and 20% as Black. The majority resided in metropolitan areas (93.3%). Mean mJOA score was 14.1 (SD 2.7, n=14). Patient interviews yielded two themes (Table 6, Figure 1).

**Table 6.** Patient Interview Themes and Representative Quotes.

| Themes | Representative Quotes |
| --- | --- |
| Experience with CSM | “It’s numbness... I can only stay asleep for maybe five hours on a good night before I have to start turning around over and over.”<br>“My handwriting has been—everyone describes it as beautiful... but anymore, it’s very jerky, sometimes illegible. It’s very frustrating.”<br>“I just take mental notes... Notice if anything’s getting better or not.”<br>“If it’s something that I need to question the doctor about, I’ll put ‘em in my notes for sure.” |
| Response to SynapTrack | “The simpler it is, the better it is I think.”<br>“That would be helpful—email, notification, or text on days I would be expected to use it.”<br>“There was a graph that showed the day, how I stacked up during the week, the month.”<br>“I would like to be able to see how I have done over time.”<br>“It would be nice to compare to myself, don’t want to know or don’t care about others.” |

*CSM = Cervical Spondylotic Myelopathy.*

### Theme 1: Experiences with CSM

Most patients relied on informal methods to track their condition:

> *“I just take mental notes… Notice if anything’s getting better or not.”*

Activities such as walking, cleaning, and sleeping served as informal indicators of symptom severity. One patient reported using a notes app to record symptoms:

> *“If it’s something that I need to question the doctor about, I’ll put ‘em in my notes for sure.”*

### Theme 2: Response to SynapTrack

All but one patient reported willingness to incorporate SynapTrack into their daily routine, citing only minor barriers such as travel disruption or lacking a waist strap for gait tasks. The most common proposed usage was at home in the morning. Nearly all patients agreed that a robust notification system would help maintain adherence:

> *“That would be helpful—email, notification, or text on days I would be expected to use it.”*

Patients provided specific feedback on SynapTrack task design, including preferences for larger instructions, simplified drawing tasks, and clear guidance on whether to prioritize speed or accuracy. This feedback directly informed iterative design refinements. Patients were neutral to positive about gamification elements but highly valued simplicity:

> *“The simpler it is, the better it is I think.”*

Many patients reported using health apps (Apple Health, MyFitnessPal, Fitbit) for step tracking and valued features showing trends over time:

> *“There was a graph that showed the day, how I stacked up during the week, the month.”*

All patients interested in using SynapTrack expressed strong desire for self-progress tracking but not comparison with other users.

### Validation of Thematic Analysis

For provider interviews, initial validation of affinity mapping categories with a third reviewer yielded 66.2% exact agreement (Cohen’s κ = 0.60, moderate agreement). Following category refinement, a second external validation achieved 91.9% agreement (κ = 0.90, near-perfect agreement). Patient interview validation demonstrated 87.3% agreement (κ = 0.87, near-perfect agreement) without need for category updates.

## Discussion

This study represents the first in-depth investigation of stakeholder perspectives on mHealth technologies for CSM. Our findings reveal consensus that current clinical assessment methods are inadequate and mHealth offers substantial promise, provided implementation challenges are thoughtfully addressed.

### Limitations of Current Assessment Tools

Providers expressed profound dissatisfaction with existing assessment tools, particularly the mJOA. Despite its widespread use as the primary outcome measure in CSM, providers characterized the mJOA as inadequate for precision medicine, a critique consistent with documented limitations including ceiling effects, limited responsiveness to change, and reliance on subjective interpretation.^5,29^ More broadly, infrequent patient-reported measures fail to capture the nuanced functional changes essential for individualized decision-making^30^ Similar limitations have been recognized in other neurodegenerative diseases, such as PD, where current assessments lack sensitivity to early disease changes.^21^ Providers in our study articulated a clear need for longitudinal, objective measures of CSM neurological function.

### Potential Clinical Applications of mHealth in CSM

Providers emphasized borderline CSM cases would benefit most from mHealth. Mild CSM management remains challenging due to variable progression rates^31,32^ and insufficient tools for counseling patients about progression risk or optimal surgical timing. Objective monitoring could help resolve this uncertainty, particularly given emerging evidence that subtle gait and dexterity changes may precede detectable clinical progression in myelopathy.^33^

Our findings also highlight two interrelated barriers to reliable longitudinal assessment: inter-rater variability and the disconnect between clinic and real-world function. Providers described how lack of standardization undermines longitudinal tracking, a concern supported by studies demonstrating significant variability in neurological examination components.^34,35^ Simultaneously, the home environment was described as “the biggest question mark,” resonating with broader recognition that clinic-based measures often fail to predict real-world performance.^36^ mHealth could address both limitations by providing standardized, sensor-based assessments in patients’ natural environments. More than just theoretical, such benefits have been shown in PD and MS, where mHealth tools have captured functional changes not detected during routine in-clinic assessments.^37–42^

Poor reliability of patient recall emerged as another limitation addressed by mHealth. Such technology enables prospective documentation of functional status and has demonstrated greater validity than retrospective self-report.^43,44^ Our findings suggest similar benefits could accrue in CSM care, particularly for tracking subtle functional changes that patients may not consciously register.

Beyond clinical utility, psychological benefits were identified. Providers described frequent patient anxiety about disease progression. The ability to objectively track one’s own status was viewed as potentially reducing this anxiety and decreasing unnecessary contacts with clinicians. This notion aligns with evidence that digital self-monitoring improves psychological well-being, sense of control, and care engagement.^45,46^ In PD, patients using continuous telemonitoring reported enhanced satisfaction, medication effectiveness, and communication with physicians.^47^ Notably, providers valued the potential for active participation in disease management.^47,48^ For CSM patients who often experience significant uncertainty about disease trajectory,^49^ visualizing their own progress could provide meaningful psychological benefit independent of clinical utility.

Importantly, patient perspectives reinforced and extended provider-identified priorities. Patients described relying on informal strategies such as mental tracking or note-taking to monitor their symptoms and expressed strong interest in structured, longitudinal self-tracking. Many patients reported prior use of consumer health applications (e.g., step tracking, fitness monitoring) and emphasized that simple visualizations of trends over time were valuable. Notably, patients consistently prioritized simplicity and clear instructions over advanced features or gamification elements, underscoring the need for user-centered design that aligns with real-world preferences.^50,51^ While providers emphasized diagnostic precision, workflow efficiency, and surgical decision-making support, patients prioritized usability and the ability to understand their own functional trajectory over time.^52^

### Implementation Barriers and Considerations

Despite enthusiasm, important implementation barriers emerged. Notably, usability concerns were paramount. Providers mentioned that patients with fine motor impairment may struggle with smartphone interfaces. Patients echoed these concerns, with participants emphasizing the importance of large text, simplified task instructions, and unambiguous performance goals (e.g., speed versus accuracy). Research in PD confirms that accommodating motor limitations through larger touch targets and tolerance for imprecise inputs can achieve adherence rates exceeding 96%,^53^ suggesting similar design principles could address usability barriers in CSM.

Equity concerns also emerged prominently. Providers recognized that socioeconomic barriers could exclude vulnerable populations. This mirrors broader digital health equity concerns that have recently intensified.^54,55^ Studies in PD found that older age groups showed more hesitancy toward digital health; however, providing instruction on device use and feedback on collected data significantly increased willingness to adopt.^56^ Such strategies warrant attention in CSM implementation efforts.

At the provider level, workflow integration emerged as a critical determinant of mHealth adoption. Providers emphasized the value of EHR integration, with the prospect of transfering findings into clinical documentation being viewed as prohibitive. Similarly, providers highlighted the importance of automated data synthesis and clinically meaningful visualization. In this context, mHealth monitoring of CSM was compared to hemoglobin A1C tracking in diabetes. Just as longitudinal A1C data have transformed diabetes management and patient engagement,^57^ providers perceived similar potential for mHealth-derived metrics to inform CSM care.

Collectively, these findings align with the Technology Acceptance Model, which identifies usefulness and ease of use as primary determinants of adoption,^58^ as well as literature demonstrating workflow compatibility as one of the strongest predictors of health technology adoption.^59^ This tension between recognized value and implementation difficulty is evident in neurology, where 73% believed remote monitoring would benefit their service, yet adoption remains limited by data integration challenges and time constraints.^60^

### Value of Early Stakeholder Engagement

This qualitative investigation employed thematic analysis to systematically uncover patient and clinician needs that will enable the creation of diagnostic and monitoring tools with the potential to integrate into patients’ lives and clinicians’ work practices. The robustness of these insights is supported by high inter-rater agreement during thematic validation. This approach demonstrates how early stakeholder engagement can inform the development of clinically meaningful digital health tools and may serve as a model for mHealth innovation in neurosurgery.

### Limitations and Future Directions

This study’s modest sample size and single-center patient recruitment may limit generalizability and nuanced perspective capture. Provider perspectives may overrepresent academic practitioners. Moreover, the study captured attitudes and preferences rather than actual behavior, and stated willingness to adopt technology may not translate to real-world engagement. Finally, these interviews were conducted early in SynapTrack’s development to inform its creation, and later stage evaluations may reveal additional findings. Future research should include longitudinal trials evaluating usability, adherence, and clinical validation, and ultimately, clinical impact across diverse practice settings.

## Conclusions

This study revealed substantial unmet needs for objective, standardized assessment tools in CSM care and demonstrated strong stakeholder interest in mHealth solutions. mHealth may bridge gaps in real-world functional assessments, reduce inter-rater variability and reliance on patient recall, support surgical decision-making, and provide psychological reassurance to patients. However, successful implementation requires careful attention to potential barriers, including intuitive user-centered design, seamless EMR integration, automated data synthesis providing actionable insights, and deliberate strategies addressing equity and accessibility. By engaging stakeholders early in development, mHealth technologies can be optimized for both clinical utility and user acceptance.

## Data Availability

All data produced in the present work are contained in the manuscript

## Acknowledgments

The authors thank the full development and testing team that has worked on the SynapTrack application.

## Notes

### Competing Interest Statement

The authors have declared no competing interest.

### Funding Statement

This study was funded by the University of Missouri Spinal Cord Injury/Disease Research Program and Washington University in St. Louis Here and Next Grant.

### Author Declarations

Institutional Review Board of Washington University in St. Louis School of Medicine gave ethical approval for this work

### Summary of Updates

Figure 1 revised to be more clear

